# COVID-19 Aerosolized Viral Loads, Environment, Ventilation, Masks, Exposure Time, Severity, And Immune Response: A Pragmatic Guide Of Estimates

**DOI:** 10.1101/2020.10.03.20206110

**Authors:** David E. Epperly, Kristopher R. Rinehart, David N. Caney

**Affiliations:** ReallyCorrect.com; California Mobile Physicians; Tahoe Blue Ltd

## Abstract

It can be shown that over 94% of COVID-19 superspreading events occurred in limited ventilation areas suggesting aerosolized transmission is a strong contributor to COVID-19 infections.

This study helps answer “How long may a person safely remain within various environments?” And “What exposure levels could result in immunity without becoming ill via asymptomatic graduated inoculation?”

COVID-19 infection likelihood, symptom severity, and immune response dependencies include viral load exposure amount. A better understanding of these relationships could help determine what Non-Pharmaceutical Interventions (NPI) would help reduce severe case counts and improve at-large epidemiologic responses in specific scenarios.

This study references peer reviewed and published studies and uses them as data sources for an estimation model that calculates infection likelihood given exposure within several example scenarios. Information from ASHRAE office ventilation standards, typical home ventilation characteristics, and an outdoor air setting are used to establish several specific examples of indoor and outdoor scenarios.

The model establishes a reference scenario using objectively measured air sample viral load concentration levels found within a carefully documented hospital environment containing 2 sick patients. The model extrapolates the reference scenario into several example scenarios that have varied exposure time duration, ventilation amount, with/without surgical mask use, activity/respiration levels, and infected subject shedding levels. It uses the reference data and scenario extrapolations to calculate an estimate of total viral load exposure dose for each scenario.

The study then interprets the various scenario total exposure dose estimates using an National Institute of Health human challenge study where volunteers were exposed to multiple specific viral quantities and observed in a clinical environment to objectively determine likelihood of infection, severity level, and immune response given each specific exposure dose. To simplify pragmatic use of the results, each example scenario presents the estimated total exposure dose alongside an intuitive severity category of *Not Ill, Minor Illness, Clinical Mild Illness, and Possible Severe Illness* which are based on a defined interpretation of the NIH study results. Immune response data related to these categories is also provided along with discussion related to asymptomatic infection, graduated inoculation, and immunity.

When appropriately interpreted for individualized applications, the estimates herein could contribute to guidance for those at low-risk for a severe case that have no obvious COVID-19 co-morbidities, with the understanding that those at higher risk should seek to avoid all exposure risk. The estimates herein may help efforts to strike a balance in developing holistic epidemiologic interventions that consider the effects of these interventions on economic, civic, social, and mental health, which have pathologies within their own realms.

## Aerosolized Transmission Premise

“A database by Gwenan Knight and colleagues at the London School of Hygiene & Tropical Medicine (LSHTM)”^4,5^ shows that over 94% of COVID-19 superspreading events occurred in limited ventilation areas suggesting aerosolized transmission is a strong contributor to COVID-19 infections. “Professors and Ph.D. students at the London School of Hygiene & Tropical Medicine have pulled together an incredibly useful database of known superspreader events from around the world. It pulls together four existing databases into one.”^6,7^

It can be seen from the London Medicine google docs spreadsheet that, out of a total of 1576 superspreading event entries, 1493 (94.7%) were classified as “Indoor”, 63 were “Indoor/Outdoor”, 4 were “Outdoor”, and the remainder were “unknown”. The large number of total cases in many of these indoor settings and anecdotal evidence of subject and index patient locations^8,9^ suggests high likelihood of aerosolized transmission and tends to rule-out “droplet or fomite” only transmission. Some anecdotal descriptive notes from the spreadsheet included: “*Every* resident in this nursing home was infected. A defective ventilation system is one of the possible causes.”, “Squash game: suspected aerosol (players used the same squash hall after each other but had no physical contact) or surface transmission”.

## Methods, Analysis Components, Data Sources, And Limitations

This study estimates the time necessary to cause various viral load exposure levels and resulting infection potential in various indoor and outdoor settings of both Influenza A and COVID-19. It uses models and math to process the results from the reference studies to calculate these estimates.

Key information needed to perform this aerosolized viral load exposure time estimation analysis and clinical severity likelihood interpretation includes ***measured amounts of SARS-CoV-2 in the air of a hospital room with COVID-19 patients*** and ***known Influenza A challenge escalation doses***; both of which are obtained from peer-reviewed and published studies. Information from ASHRAE Office Ventilation standards and an Outdoor Air Exchange model are also utilized. This section also covers limitations of the various analysis components as an integral part of their detailed explanations.

### Analysis Components Overview

The analysis in this study requires various “known” data sources that are fed into a mathematical “spreadsheet” formula that calculates estimations of the time necessary to cause various viral load exposure levels in various environments. These exposure levels are then interpreted into “symptom level” categories such as *“Not Ill”, “Minor Illness”, “Mild Illness”, and “Possible Severe Illness”*.

To perform this analysis, the following “data sources” and “analysis model components” are required:

1. Aerosolization Model (provided herein)
2. Outdoor Air Exchange Model (provided herein)
3. Known Challenge Dose Escalation Results Data for Influenza A (challenge dose for SARS-CoV-2 is not yet known)^10^
4. Measured SARS-CoV-2 aerosol concentration in a known environment containing infected patients^11^
5. Common Respiration Rates (selected example rates from references provided herein)
6. General Office Building Ventilation Standards^12^
7. Influenza A Versus COVID-19 Infectivity Adjustment (data extrapolation from references provided herein)
8. Pre-symptomatic vs Hospitalized Viral Loads (data extrapolation from references provided herein)

The details of these data sources, analysis model components, and related limitations are elucidated the sections that follow.

### Aerosolization Model

A virally infected person will generate aerosolized virions and it has been demonstrated that significant COVID-19 transmission modes include aerosolized virus within indoor settings^13,14^. The aerosol fluid is largely water with amounts of mucosal secretions and viral elements^15^. This mostly water-based aerosol diffusion is complex and depends upon factors including temperature, humidity, barometric pressure, air exchange rates, turbulence, aerosol generation rates, etc. When comparing the effect of different air exchange rates on a given air viral concentration relative to a reference air exchange rate, a simple linear calculation is used for purposes of estimation within this study. This linear calculation provides approximate estimates which can provide pragmatic benefit. A better aerosolization model would likely be helpful and could be part of further work, although it may complicate the desire for “easily understood and actionable rules of thumb” regarding NPI safety protocols. Improving the accuracy without complicating the presentation of estimates would be a good goal for further work.

SARS-CoV-2 and Influenza viruses are both enveloped, single-stranded RNA viruses, and both are encapsidated by nucleoprotein and have enough similarity that they can be compared for purposes of aerosolized transmission characteristics.^16^

Since this study considers individuals that are more than 2 meters distant from the infected individuals, large droplet and surface contact conditions are not considered. Those considerations must be accommodated and estimated separately when they apply. Other safety protocols such as hand-washing, surface-disinfectant, droplet avoidance, etc. are well defined in other studies and resources. This study seeks to add safety protocol information regarding exposure times to infectious aerosols in various environments.

### Outdoor Air Exchange Model

In the following diagram, a 15×15×15 foot cube (3375 cubic feet) of air is moved 15 feet to the right by a 3 MPH wind (264 feet per minute). **This constitutes 1 complete air exchange of that 3375 ft**^**3**^ **volume in 0**.**057 minutes**^**17**^ **(1 air exchange every 3**.**4 seconds, 17**.**6 air exchanges per minute, 1056 air exchanges per hour) which is the equivalent of 59**,**400 cubic feet per minute (CFM)**.

This simplified laminar air flow model establishes the effect of a minimal / low terrestrial wind condition such as 3 MPH as part of a slightly conservative approach to modeling an outdoor environment as majority prevailing wind scenarios are often larger than 3 MPH. An environment with lower wind conditions and/or obstructions would differ as would higher wind conditions than this example “open air” model that serves as a specific example that functions as an example pragmatic use-case. It is noteworthy that this single 3 MPH example calculated to such long periods of relative safety that this single example provides a pragmatically useful outdoor example.

### Known Challenge Dose Escalation Results

An NIH study entitled ***Validation of the Wild-type Influenza A Human Challenge Model H1N1pdMIST: An A(H1N1) pdm09 Dose-Finding Investigational New Drug Study (*Memoli MJ, *et-al)***^***18***^ demonstrated that low viral load exposures caused people to develop increased immunity levels without becoming significantly sick – they “seroconverted while having minimal clinical illness and no shedding”.

It can be seen from the Memoli study that symptoms increased significantly at 10^5 TCID50 exposure in an influenza A (H1N1 challenge study. Shedding significantly increased at 10^6. Another human challenge study found similar results in “A Dose-finding Study of a Wild-type Influenza A(H3N2) Virus in a Healthy Volunteer Human Challenge Model”^19^

**Throughout this study, the “Dose” listed above will be associated with symptom categorizations as follows:**

- **10^3 = *<u>Not</u> <u>Ill</u>*: none with MMID, majority no symptoms, no shedding**
- **10^4 = *<u>Minor</u> <u>Illness</u>*: none with MMID, half with symptoms, no shedding**
- **10^5 = *<u>Mild</u> <u>Illness</u>*: 20% with MMID, majority symptoms, some shedding**
- **10^6 = *<u>Possible</u> <u>Severe</u> <u>Illness</u>*: majority with MMID, majority symptoms, about half shedding**

This study considers that the common use of the phrase “COVID-19 Mild Case” best matches the symptoms of “mild to moderate influenza disease” (MMID) in terms of the illness experience. Anything without MMID would not be “Flu like”, but might be cold or sore-throat like and this study considers that “*Not Ill*” or “*Minor Illness*” given the severity concerns surrounding COVID-19 cases. The “*Not Ill*” characterization is more likely to have no symptoms per the Memoli study.

Since 20% of those in the Memoli study challenge group experienced MMID at the 10^5 exposure level, this was probably the best exposure level to use to describe “*Mild Illness*” / “COVID-19 Mild Case”.

Since the majority receiving 10^6 and above exposure levels experienced MMID, this is considered the level at which “*Possible Severe Illness*” might develop – conversely, below this level, development of severe illness necessitating medical attention would be expected to be extremely rare. Since COVID-19 and Influenza A severity have many determinants, these categorizations assume good general health and no significant co-morbidities. While these “power of 10” boundaries are only approximately reflective of reality, they are helpful for estimation.

***These symptom categorizations and this study does NOT apply to skilled-nursing patients, obese (BMI > 30), or those with cardiac, immune, cancer complications, etc***.

The Memoli study excluded BMI > 40. A BMI > 30 constraint was chosen for this study to better accommodate the conventional understandings of COVID-19 co-morbid risk factors and erring on the side of caution when considering this study’s estimates. Basing illness interpretations upon Memoli’s small N is supportable when considering the multiple order of magnitude graduated dose increases below the MMID threshold and subject co-morbidity screens (cardiac, BMI, etc.) applied herein. Application of the illness interpretations should consider that small N.

### Measured SARS-CoV-2 Aerosol Density In A Known Environment

Per **“Viable SARS-CoV-2 in the air of a hospital room with COVID-19 patients” (Lednicky JA, et-al)**^20^, it was found that “viable virus was isolated from air samples collected 2 to 4.8m away from the [two] patients” in a 3.5 x 7.0 meter hospital room with **“six air changes per hour”** (0.1 air exchanges per minute). “Estimates of viable viral concentrations ranged from 6 to 74 TCID50 units/L of air.”. **Regardless of the number of infected patients, this study uses the mean viral concentration of 40 TCID50 units/L of air as the reference environment that contains one or more sick persons**.

Since the ceiling height was not specified, it can be estimated from “Ceilings in patient bed areas including Bed Rooms, Bed Bays and Recovery areas should be a minimum of 2700mm.”^21^ Therefore, the entire room can be estimated to be **66 cubic meters (2331 cubic feet). Normalized to the Outdoor Air Exchange Model, this would be a room of 3375 cubic feet at 337**.**5 CFM producing 6 air exchanges per hour (0**.**1 air exchanges per minute)**.

No specific mention of patient face masks was noted in the reference hospital room, so this study assumes none were in place on these patients. It appears that at least 1 of the patients was strongly positive with a 32 Ct PCR test, but these 2 hospital patients may not be among the strongest level of shedding patients. They may or may not represent a “typical” hospital patient shedding scenario.

This study uses a “perfect mixing steady state” static equilibrium aerosol diffusion and dilution approximation model that scales linearly with air exchange^22^. For example, given a reference scenario of a specific stable aerosol concentration at 6 air exchanges per hour, that concentration would approximately diminish to half of the reference concentration at 12 ACPH.

This study’s model of air exchanges does not consider area population densities such as “cubic feet of available air per person”. Perhaps obviously, a large warehouse with 2 people at 6 air exchanges per hour would likely result in lower aerosol concentrations than a small room with the same 6 ACPH. This study uses a hospital room with specified dimensions as the reference case, so population density adjustments would need to be made for crowded establishments, sparsely populated warehouses, etc. Diffusion and dilution realities are more complex than the model used in this study and better estimates could be achieved with better models.

It would be preferred to have more data regarding indoor aerosolized viral concentrations at distances of approximately 2 to 5 meters from an infected individual. Having only 1 reference environment is a result of a time limitation to perform this study. Adding additional viral load concentration reference points to Table 2 would be desirable.

**Table 1.** Dose Escalation Results.

| Dose | Male | Female | Total | Symptoms | Viral Shedding | Both (MMID) | 4-Fold Rise in HAI Titer |
| --- | --- | --- | --- | --- | --- | --- | --- |
| 10<sup>3</sup> TCID<sub>50</sub> | 3 | 2 | 5 | 2 (40%) | 0 | 0 | 1 (20%) |
| 10<sup>4</sup> TCID<sub>50</sub> | 4 | 0 | 4 | 2 (50%) | 0 | 0 | 0 |
| 10<sup>5</sup> TCID<sub>50</sub> | 2 | 3 | 5 | 4 (80%) | 1 (20%) | 1 (20%) | 1 (20%) |
| 10<sup>6</sup> TCID<sub>50</sub> | 10 | 9 | 19 | 17 (89%) | 9 (47%) | 9 (47%) | 16 (84%) |
| 10<sup>7</sup> TCID<sub>50</sub> | 9 | 4 | 13 | 11 (85%) | 10 (77%) | 9 (69%) | 11 (85%) |
| Total | 28 | 18 | 46 | 36 (78%) | 20 (43%) | 19 (41%) | 29 (63%) |

**Abbreviations:** MMID, mild to moderate influenza disease; TCID<sub>50</sub>, 50% tissue culture infectious dose.

**Table 2.**
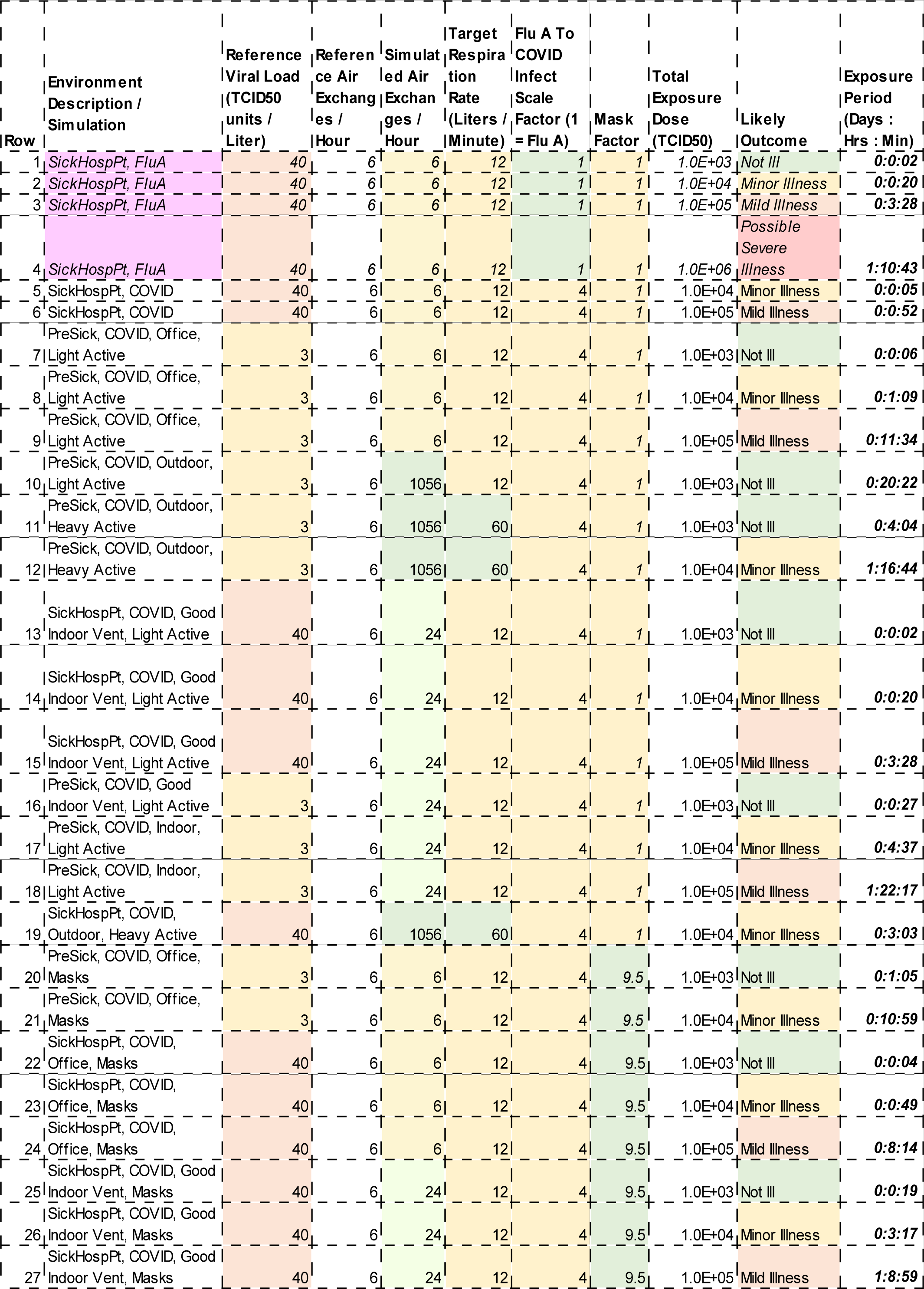
shows the multiple potential scenarios based on a spreadsheet formula^32^.

### Respiration Rates

Adult respiration rate “while resting is about 5–8 litres per minute”, and “light activities minute volume may be around 12 litres”^23^ and “moderate exercise may be between 40 and 60 litres per minute”. In this study, the **respiration rates of 12 L/m and 60L/m will be considered for the person being exposed**.

### General Office Building Ventilation Standards

“According to ASHRAE Standard 62.1, an office will require 4-10 air changes per hour depending on the occupancy and size of the office.”^24^ In an example ventilation “rule of thumb” example, **a room of 6912 cubic feet having 6 air exchanges per hour would require 691 CFM**.^25^

**Normalized to the Outdoor Air Exchange Model, this would be a room of 3375 cubic feet at 337**.**5 CFM producing 6 air exchanges per hour (0**.**1 air exchanges per minute)**.

### Influenza A Versus COVID-19 Infectivity Adjustment (Multiplier)

Since no viral load challenge data for COVID-19 could be found at time of writing, this study must adjust estimates from challenge data for Influenza A to estimate SARS-CoV-2 infectivity.

SARS-CoV-2 has been claimed to have R0 values ranging from 1.5 to 3.5 to 5.7^26^ depending upon estimation sources. The mean of these 3 numbers is about 3.6. H1N1 Influenza R0 is widely estimated to be near 1.46 to 1.48 with a mean of 1.47.^27^ The R0 means of Influenza A vs COVID-19 R0 calculate to a ratio of about 1 to 2.5.

Using Infection Fatality Rates (IFR) of 0.1% for Influenza A and 0.4% for COVID-19 shows about a 1 to 4 ratio.

Total Fatalities for a typical Influenza A year vs COVID-19 appear to be about a 1 to 3 ratio as of September 2020.

The mean of these 3 ratios calculates to 3.2. **This study chose to round up to 4 with a bias toward representing COVID-19 as much more aggressive than Influenza A**, which creates more “conservative” estimates that result in additional “safety margin”. The reader / observer is free to re-calculate the spreadsheet times based on a different “Influenza A To COVID-19 Infectivity Multiplier” as this adjustment multiplier value accuracy is a limitation of this study and is admittedly weakly supported. This adjustment value and/or it’s model dependencies could be updated when if/when human challenge data for SARS-CoV-2 becomes available.

Based on the aerosolization model defined above, this study considers that viral shedding levels of both Influenza A and COVID-19 are similar for patients of similar infection severity. Thus, from the shedding perspective, Influenza A and COVID-19 shedding data are considered equivalent in this study in the sense that the viral load concentrations as a result of various aerosolization mechanisms abide by fundamentally similar aerosol diffusion physics (e.g. a person with a given PCR Ct breathing normally will shed same viral load independent of Influenza A or SARS-CoV-2 virus type). The physics of aerosolization are considered identical for Influenza A and SARS-CoV-2 as the enveloped viral sizes and physical characteristics within aerosols are virtually identical. The Infectivity Adjustment accommodates the apparently significant differences between in-vivo infectivity of a given viral load concentration of the different virus types.

### Pre-symptomatic vs Hospitalized Viral Loads

It has been shown in various challenge studies as noted earlier that case severity tends to increase with increasing initial exposure viral load. Patient viral loads also vary dramatically depending upon stage of illness.

Per ***Validation of the Wild-type Influenza A Human Challenge Model H1N1pdMIST: An A(H1N1) pdm09 Dose-Finding Investigational New Drug Study***^***28***^ ***Figure 1***, it can be seen that symptoms typically became observable within 3 days of challenge. From data in that study relating to the pre-symptomatic phase, the viral load contributing to shedding at day 1.5 was about 10^3 and was 10^4 or higher during the peak symptom period. This represents a 10 fold increase between the median pre-symptomatic time period and the peak symptom period, keeping in mind that patients in this study did not experience severe symptoms that would require hospitalization, though they were studied in a clinical environment.

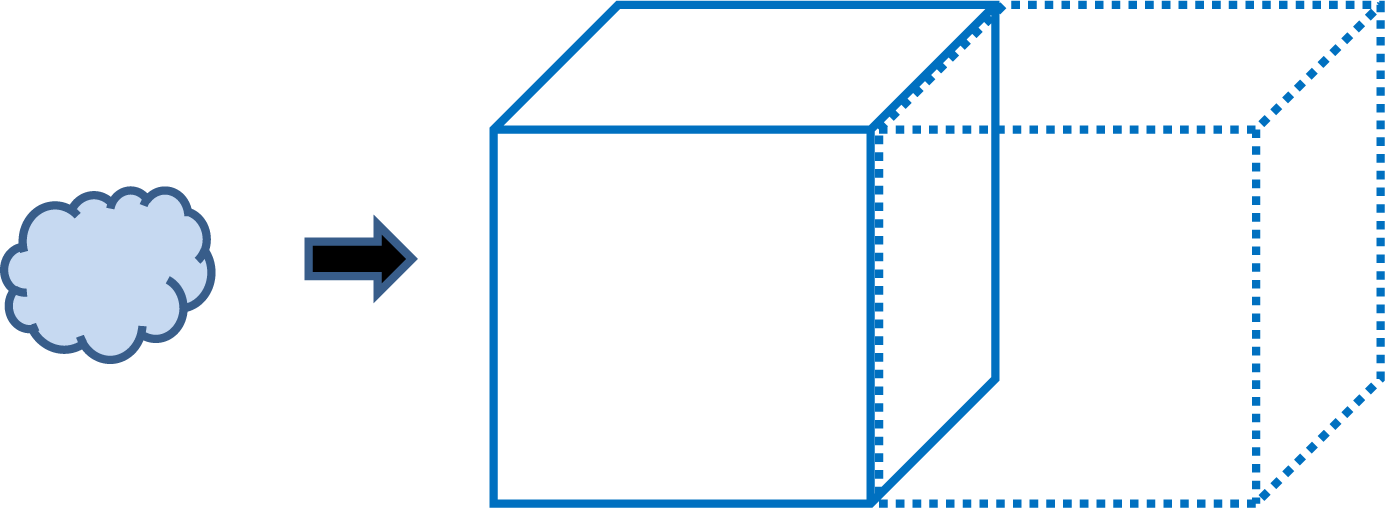

Per “Viable SARS-CoV-2 in the air of a hospital room with COVID-19 patients”^29^ it is known that both patients were hospitalized which would suggest that their cases were more severe than a typical mild COVID-19 case and that maximum shedding would be greater than mid-phase pre-symptomatic shedding for these same patients. Patient 1 had a PCR Cq [similar to Ct] of 32 which is 2^4 (16x) less than that of a typical Cq positive threshold of 36 or below (some use Cq < 40)^30^. Patient 2 had an unspecified Cq. The hospitalized COVID-19 patients experiencing a more severe case COVID-19 than that of the Influenza A study suggests that the patients tend to have longer periods of symptoms and shedding than that of the Influenza A study.

Taking the average of the 10 fold increase from the Influenza study and a 16x Cq from positive threshold in the COVID-19 hospital air study, this study uses a roughly 13 to 1 ratio of symptomatic to pre-symptomatic shedding multiplier. That is to say that during mid-point of pre-symptomatic period (median of time to symptoms), the shedding is about 1/13^th^ that of a hospitalized COVID-19 patient during the mid point of their hospitalization. **So the air viral load was adjusted from 40 TCID50 units / liter as measured in the hospitalized COVID-19 patients to an estimate of 3 TCID50 units / liter for the mid-phase pre-symptomatic individual modeled in this study**. Due to rapidly rising viral loads during the pre-symptomatic phase of infection, any estimate is widely dependent upon the viral load replicating within the infected individual. When considering *Exposure Period* estimates in Table 2, the likelihood of encountering someone who is already experiencing minor symptoms, or nearly so on the illness timeline, should be considered.

### Mask Usage

A study on mask efficacy suggests that the Exposure Period estimate in the table could be adjusted to accommodate mask usage.

“Surgical masks nearly eliminated viral RNA detection in the coarse aerosol fraction with a 25 fold reduction in the number of viral copies, a statistically significant 2.8 fold reduction in copies detected in the fine aerosol fraction, and an overall statistically significant 3.4 fold reduction of viral copy number in the exhaled aerosols.”^31^

Given the aerosol focus of this study, aerosol data from this study is provided and discussed so that the study Exposure Period estimate may be adjusted. **If the “infected” person wears a mask, the estimate could be multiplied by 3**.**4. If the “infection target” person wears a mask, the estimate could be multiplied by 2**.**8. To have the estimation reflect both “infected” and “infection target” wearing masks, the estimate could be multiplied by 9**.**5**. Additional mask study data would improve the estimates in this study.

## Results

**Comparing a 3375 cubic foot office with 6 air exchanges per hour (per ASHRAE Standard 62**.**1) with an comparable sized open space outdoor setting with a 3 MPH breeze (1056 air exchanges per hour) shows that a similarly sized outdoor space has 176 times more air exchanges over any given time period than a small office or a hospital room similar to the reference cases used in this study**. An open outdoor space is further superior due to its virtual “infinite ceiling”. Additionally, indoor spaces tend to have areas of stagnant air flow around the cubicles, furniture, and other semi-enclosed spaces within the room.

Table 2 considers the scenario of non-infected person[s] in an indoor environment within 2 to 4.8 meters of two COVID-19 infected persons and estimates the Exposure Period related to *Likely Outcome*s. The entire table uses shedding equivalency for Influenza A and COVID-19 as stated previously (aerosol physics is identical). Rows 1 through 10 consider the case of non-infected person[s] in the environment performing light activity functions and respirating accordingly, while rows 11 and 12 consider a person engaging in heavy exercise or otherwise respirating at 60 liters per minute.

Rows 1 through 4 of Table 2 provide documentation and analysis of the “reference” environment which has 2 hospitalized COVID-19 patients. It establishes 4 total exposure dose levels using the Memoli Influenza A study and uses an Influenza A infectivity model. From those 4 rows, it can be seen that the non-infected visitor not wearing any PPE / masks would likely remain *Not Ill* when present for less than 2 minutes, and perhaps develop a *Minor Illness* after 20 minutes, and likely having *Mild Illness* after 3.5 hours, and almost assuredly becoming Ill after 34 hours. Immune response and some level of immunity could begin in as short as 2 minutes, but more likely would occur after more than 3.5 hours. The visitor would be unlikely to shed any virus until around or after a 3.5 hour visit. Again, this considers patients ill with Influenza A using direct Influenza A Challenge Study data.

Rows 5 and 6 use estimations to model the 2 hospitalized COVID-19 patients with visitor result using a COVID-19 infectivity model. From those 2 rows, it can be seen that the non-infected visitor not wearing any PPE / masks would perhaps develop a *Minor Illness* after 5 minutes, and likely experiencing *Mild Illness* after 52 minutes in a 6 ACPH environment.

Rows 7 through 12 use estimations to model the result as if a pre-symptomatic person was infected with COVID-19 but NOT yet far enough into the infection as to have symptoms and large amounts of aerosolized shedding that would likely be present in a hospitalized patient with symptoms requiring hospital admission. In rows 7 through 9, it can be seen that the non-infected visitor would perhaps develop a *Minor Illness* after 1 hour, and likely experiencing *Mild Illness* after 11 hours.

In row 10, the estimation is adjusted to simulate air exchange of an outdoor environment with a 3 MPH wind as per description in *Methods*. It can be seen that the non-infected visitor not wearing any PPE / masks within 2 to 4.8 meters of 2 infected persons would likely remain *Not Ill* when present for less than 20 hours. This 20 hours estimate clearly shows the contrast between indoor environments that have no special ventilation measures and outdoor settings. Indoor settings with strong ventilation improvements could begin to approach those of an outdoor setting.

In rows 11 and 12, the estimation is adjusted to simulate the same outdoor environment, but with the non-infected visitor exercising around 2 resting pre-symptomatics. It can be seen that a non-infected person would likely remain *Not Ill* when present for less than 4 hours and perhaps develop a *Minor Illness* after 40 hours. This implies that outdoors is low risk even without masks. While it was considered too complex to estimate the case of 2 pre-symptomatics exercising heavily with an uninfected person outdoors, it may be possible extrapolate this scenario from the table.

In rows 13 through 18, the estimation is adjusted to simulate an indoor environment with 24 air exchanges per hour, which is 4 time more ventilation than the reference office. Similar to the previous rows, individuals ill with COVID-19 and pre-symptomatic individuals are considered. In row 19, an outdoor environment is simulated with a person exercising near individuals ill with COVID-19. Rows 20 through 27 add surgical mask considerations. An N-95 mask would have even better performance.

## Discussion

From Results, it can be seen that *Likely Outcome*s vary widely based on ventilation conditions and level of shedding of the infected individual. While the estimates predict that most outdoor settings with as low as a mild 3 MPH breeze are likely to result in no or *Minor Illness*, there are many determinants associated with severe cases including co-morbidities such as skilled-nursing-facility resident status, cardiac health issues, obesity, cancer, immune-compromised, etc. The “*Results Estimates*” assume good general health and no significant co-morbidities as stated earlier.

In Table 2, the “*Likely Outcome*” column study calculates for the scenario where there are known to be a few infected within any given social setting. In a real population that has, for example a 100 per 100,000 population infection-rate, that “*Likely Outcome*” would have a 1 in 1000 chance of occurring. In the other 999 of 1000 cases, since no-one is infected, no-one can become infected, so the result is *Not Ill*. This study does NOT consider that 1 in 1000 probability that lowers the probability of becoming ill in a “real life scenario”. The estimates in this study consider the case of any number of healthy individuals being within 2 to 4.8 meters of 2 definitely infected individuals (the hospital reference case used as the baseline for all estimates) so that a “worst case probability” scenario can be estimated. The probability of infection could be computed separately based on “local infection rates” and then further adjusted based on the Illness “*Likely Outcome*” for any given exposure period.

All estimates are subject to prior mentioned limitations. All times are specified as Days:Hours:Minutes. The term “Office” implies a room with 6 air exchanges per hour (ACPH) as Table 2 defines unless otherwise noted. All other aspects can be also observed in Table 2 and prior commentary. As mentioned in the “Mask Usage” section, the term “mask” refers to a surgical mask.

From Table 2, many personal and business scenarios can be considered. The border between “*Not Ill*” and “*Minor Illness*” can help determine risk profile for virtually any scenario. The term “*Mild Illness*” is a clinical definition, and for COVID-19 can mean quite unpleasant flu-like symptoms including fatigue, fever, chills, muscle aches, headache, sore-throat, etc. that would not require hospitalization. The COVID-19 definitions of Moderate and Severe mean they most often require clinical attention and/or hospitalization. “*Minor Illness*” can mean “common-cold” like symptoms or slightly worse but not full “Flu Like” symptoms which is defined by “*Mild Illness*”. The COVID-19 Moderate and Severe cases are referred to in Table 2 as “*Possible Severe Illness*”. See the “*Known Challenge Dose*” section for more detail. To help clarify Table 2, here are a few observations that might be made:

A person wearing a mask doing light work (*Light Active*) near a COVID-19 hospital patient (*SickHospPt, COVID*) wearing a surgical mask in a 6 ACPH patient room may encounter enough exposure to develop a *Minor Illness* in a 49 minute period (0:00:49); but would likely not develop *Mild Illness* “flu like” symptoms unless in that environment for over 8 hours (0:08:14). Using N-95 and other improved PPE would have the effect of increasing those estimates. It is possible that immunity may begin developing at even shorter intervals. It is possible that by rotating medical staff work through COVID-19 patient areas at exposure intervals below the low end of this range several days apart and monitoring antibody test levels could help provide a path to immunity and increased ability to work with increasing exposure level environments without illness. Encountering short to increasing length exposures that result in graduated inoculation and avoiding lengthy exposures until antibody test titers become strongly positive may be a path to avoiding healthcare staff illness. It is likely several short-term exposures separated by several days will increase antibody test titers, and as they reach protective thresholds, longer exposures would likely increase those antibody test titers to a level of sterilizing immunity. Repeat exposure would likely maintain that sterilizing immunity indefinitely. Having non-immune medical staff avoid lengthy exposure to patients that have high shedding potential based on PCR Ct counts could also help avoid medical staff illness. It is possible that some anecdotal observations of lower rates of medical staff infection over time correlates to unplanned graduated inoculation. Better planning as elucidated herein could provide better results.

People wearing masks doing light work in a typical 6 ACPH office with a pre-symptomatic individual nearby may encounter enough exposure to develop a *Minor Illness* that is “cold like” in around 11 hours (0:10:59).

People wearing masks doing light work in a typical 6 ACPH office with a sick individual nearby may encounter enough exposure to develop a *Mild Illness* “flu like” in 0:08:14, meaning that with masks, an 8 hour shift near a sick person, both wearing masks, would likely result in those within a few meters becoming infected. With better ventilation at 24 ACPH, this would likely extend to more than 32 hours (1:08:59).

People not wearing masks doing light work in a 3 MPH open area outdoor breeze with a pre-symptomatic individual nearby would likely not encounter enough exposure to become ill until after over 20 hours (0:20:22). People not wearing masks doing heavy exercise in a 3 MPH open area outdoor wind with a sick individual nearby may encounter enough exposure to develop a *Minor Illness* “cold like” in around 3 hours (0:03:03). People not wearing masks in a typical 6 ACPH office with a pre-symptomatic individual nearby would remain *Not Ill* in an encounter that is less than 6 minutes. Most homes with mostly closed windows have between 0.2 and 2 ACPH^33,34^. In a home, exposure durations in Table 2 would be reduced by 3x to 30x when compared to a typical office building with 6 ACPH (e.g. row 6 might be as low as 1.7 minutes in a home).

In a hair-styling appointment scenario, if done within a common 6 ACPH indoor environment with no-one wearing masks and a pre-symptomatic person, *Minor Illness* might occur in around 1 hour (0:01:09), whereas that would extend to (0:04:37) in a 24 ACPH environment, which would significantly reduce the chance of illness in a 1 hour appointment.

Wearing masks would further improve the likelihood of being *Not Ill*. In the 24 ACPH environment with masks even with a sick person, the other person would likely remain *Not Ill* after 19 minutes (0:0:19). In the scenario of a 2 hour movie at a theatre in a 24 ACPH environment without masks and with pre-symptomatic persons, *Minor Illness* might occur in around 4 hours (0:04:37). If masks were used, even if a sick person were present, *Minor Illness* might occur in 0:03:17.

Not wearing a mask and being in proximity to a sick person indoors with 6 ACPH for 5 minutes could result in *Minor Illness*. Extending that to 52 minutes could result in *Mild Illness*. Passing them briefly for less than a minute would likely leave one *Not Ill*. This scales to hours and days when outdoors – certainly of low concern for brief encounters – even without a mask. Regarding brief encounters, keeping calm and moving along is usually safe even when around people who are known to be sick. For pre-symptomatics and when wearing masks, these numbers scale accordingly to even longer periods of safety – see Table 2.

It is noteworthy that once an individual has reached a target exposure level, the individual should attempt to eliminate further exposure for several days so that the adaptive immune system may respond to the exposure before encountering additional exposure. A level of exercise after exposure can improve the probability and speed of immune response by accelerating viral movement into the Secondary Lymphoid Organs’ (SLO) germinal centers.^35,36,37,38^

General immunology and the Influenza A Challenge study referenced earlier have demonstrated that a person becoming sick begins to lightly shed virus about 2 days prior to symptoms and rapidly ramps infectivity to a peak that occurs about 1 day after symptoms begin; whereupon a normal immune system responds to reduce infectivity within a few days to a week or so of symptom onset. This may depend upon general individual health and care in using optimal balances of rest and light exercise during recovery.^39^ Some youth and extreme athletes are able to knock down illness within hours or a couple of days.

By knowing the type of individual that is most likely to be encountered (pre-symptomatic or sick) and the likelihood of encountering an ill person (e.g. “1 in 1000” or “Confirmed Sick”), one may discover a time limit estimate that meets one’s personal risk tolerance. It is important to keep in mind that these are estimates and the actual times could vary widely based on a large number of factors that even include amount of sleep, eating habits, exercise, and other factors too numerous to express.

Personal, business, and public health guidelines must consider many factors including number of active infections per capita, general area population density, specific environment occupancy, temperature, respiration rates, loudness of vocal activation, general population health, etc.^40,41,42^ Table 2, the studies that feed it, and other related studies, may help determine reasonable health guidelines and be useful to the general public in setting individual risk tolerances. Businesses may benefit from having this study’s actionable quantitative estimates inform specific business decisions including ventilation enhancement opportunities. Given diverse business environmental characteristics and the limitations of this study, it is recommended that this study help inform local entity specific and personal / individual decisions. It is not intended for use in specifying government policy. For example, each of the following might select different exposure risk tolerances:

- A person in good health who rapidly and fully recovered from COVID-19 (recently recovered / immune)
- An unexposed (COVID naïve) pro / amateur athlete in excellent health
- An unexposed (COVID naïve) middle-aged person in good health

Every individual’s risk tolerance and optimal exposure level is different. Those already recovered may benefit from light re-challenges that preserve IgA IgM antibody titer levels and related protective or even sterilizing immunity^43,44,45,46^. Every business building has attributes that may favor applying different estimates.

***Importantly, those with co-morbidities that place them at high-risk for a severe case should target zero exposure. Co-habitants and care-givers who attend to these high-risk individuals should use similar precaution***.

Those who are at lower risk for a severe case and are not in contact with high-risk individuals could opt for a more “normal” lifestyle and use Table 2 to guide them selecting a conceptual boundary of “*Not Ill*” and “*Minor Illness*” in accordance with personal risk preferences. It is possible that becoming exposed to very small viral loads could result in some level of protective immunity as a result of an asymptomatic infection^47^.

Those who have already recovered from an infection could select boundaries that are more likely to help them maintain immunity without being exposed to an excessively strong (or weak) re-challenge. In healthy individuals, properly timed and dosed re-challenges may be helpful to maintain sterilizing immunity that protects those not yet infected during months-long outbreaks^48^. Estimates such as these, as accuracy is improved through future research, provide the structure for an objective data-science based methodology for avoiding illness and maximizing immune response.

These specific quantitative examples of exposure viral load versus symptoms and immune response could increase public understanding and consciousness of concepts such as “viral load”, “exposure time”, “challenge dose levels”, “shedding quantities”, “immune seroconversion”, “re-challenge”, “protective immunity”, and “sterilizing immunity”. New levels of personal hygiene that complement centuries-old adages such as “wash your hands” could become common-place.

Understanding the viral load threshold differences between protective vs pathologic exposure could help guide individual behavior. In a not-distant future, the COVID-19 era’s prevalence of mystery and fear could be replaced by scientific understanding and individualized immune system management innovation. It is epidemiologically possible to improve the probability of developing immunity without becoming ill. While the estimates herein are of insufficient accuracy for establishing that clear guidance, they may be of some utility to a portion of the population; and the concepts presented herein could be developed such that guidance of increasing reliability becomes available over time.

If primary education included content similar to this study as a health class assignment, a tremendous epidemiologically protective step forward would accompany the next generation.

### Disclosures

The authors performed all work on this study without compensation as an act of good will and statesmanship and have no beneficial affiliations with any institutions related to the work.

### Limitations

***While the estimates in this study cannot be considered authoritative due to estimation limitations, they offer considerably greater resolution and accuracy than WHO and CDC recommendations such as “6 feet of distance” and “use facial coverings” which do not differentiate between exposure periods, environments, and severity***.

The estimates in this study could be a significant improvement over current WHO and CDC guidelines in that they provide specific quantitative number estimates extrapolated from peer-reviewed studies for various environments. The various environments include typical ASHEAE office ventilation standards, one example of dramatically improved ventilation (4x), and outdoor 3 MPH conditions. The estimates also consider the infected person as either very sick or pre-symptomatic, the level of exertion of those breathing in vicinity of the infected, and various *Likely Outcome*s from *Not Ill* to *Possible Severe Illness*. It is desirable to add near-freezing temperature environments as it is predicted that these environments could solidify a viral load preventing diffusion and dilution, yet increasing precipitation.

This study considers aerosolized transmission at a distance of > 2 meters. The estimations in the table focus on aerosolized transmission and do not consider the case of droplet or fomite (surface contact) based transmission which can be averted through some distance, barriers, surface disinfection, socially hygienic behavior (not sneezing at your neighbor), hand-washing, and other Non-Pharmaceutical Interventions (NPI). Droplet scenarios tend to have short-lived exposure periods relative to aerosols as the droplets rapidly precipitate. Fomite disinfection may also reduce exposure period related risks.

While a reader may personally choose to consider the estimates for personal, business, and other guidelines, they have no established range of accuracy, assurance, guarantee, medical efficacy, or legal standing. In the absence of better information, these estimates should be considered only with additional good judgement and after further verification. Human response to pathogens is widely variant and these estimations assume the normal healthy population with better than average immune response, no co-morbidities, and a very specific set of scenarios. Any application of these estimates is at the risk of those applying them. There may be unforeseen errors in the estimation, extrapolation, and underlying study interpretation. There may be errors in the referenced studies. No liability may be assumed or implied. The primary purpose of this study is to provide a conceptual framework for further study, interpretation, and application. Choice to observe any of the specific results or commentary belong solely to the reader.

As additional data becomes available, this study could be revised to reference and include that additional data. The estimation methods used in this study could also be improved. Together, additional data and improved estimation methods could be used to revise the spreadsheet that makes the objective computations found in Table 2. Future studies could improve models and estimation accuracy. **While the values in Table 2 are specific, they cannot be relied upon as accurate as they are estimates from an approximation model**.

## Data Availability

The manuscript contains all relevant data and references.

